# Heat inactivating and reusing of virus-contaminated disposable medical mask

**DOI:** 10.1101/2020.07.01.20144527

**Authors:** Wuhui Song, Bin Pan, Haidong Kan, Yanyi Xu, Zhigang Yi

## Abstract

As the prevalence of COVID-19 in all the area of China and continuous spread to many other countries, it is in urgent need of medical mask for community people. We have reported a simple method to inactivate artificially influenza virus-contaminated medical mask without affecting the filterability of mask by heating with hair dryer for 30min. In this study, we extended our studies to optimize our protocols and evaluate the filterability of mask after several-rounds of heat treatment. We found that baking at 70°C for 30min almost completely inactivated virus as hair dryer treatment for 30min described before. One-round of heat treatment with hair dryer for 30min or baking at 70°C for 30min did not affect filterability of mask whilst two-rounds of treatment slightly but significantly reduced filterability of mask. Thus, heating with hair dryer for 30min or baking at 70°C for 30min can inactive contaminated medical mask and the treated mask can be reused at least once.

## Introduction

Since the outbreak of COVID-19 in Wuhan China, it was endemic to all over the China within less than one month, causing over 80000 cases and over 3000 mortalities in China. It is considered to have pandemic potential, as it has been spread to almost all the continents and has caused over 20000 cases outside China. COVID-19 is caused by infection with a novel coronavirus SARS-CoV-2^1-3^, and its human-to-human transmission occurs mainly through close contacts^4^. Personal protective equipment (PPE) like medical mask is critical for public to prevent the infection of COVID-19. Thus it is in urgent need of medical mask all over the world. However, there is global shortage of medical mask. Medical mask acts similarly as N95 respirators for preventing Influenza virus infection ^5^. Re-use of disposable medical mask would alleviate the panic of shortage of mask. Prior to re-use of mask, we should inactivate potential contaminated virus on the mask. Heat inaction of virus such as incubating at 56°C for 30min is widely accepted to be able to inactivate enveloped virus like influenza virus and coronavirus. And importantly, heating at 70 °C for 24 hour does not significantly change the filtration efficiency of polypropylene (PP)^6^, a core matrix for medical mask, whereas conventional disinfectant like ethanol dramatically destroys filter performance of polypropylene electret filters ^7^. We have developed a simple method to inactivate medical mask that is artificially contaminated by influenza virus via heating with hair dryer for 30min without affecting its filterability ^8^, which can be easily adapted by public. In this study, we extended our studies to optimize our protocols by shortening treatment intervals with hair dryer and baking mask at different temperatures. We demonstrated that hair dryer treatment for 15 min did not effectively inactivate influenza virus (IVA) on medical mask compared with 30 min as described before. And that baking at 70°C for 30 min effectively inactivated IVA on mask whereas baking at 65°C for 30 min did not. We also evaluated the filterability of mask after several-rounds of heat treatment. Our results reveal that two-rounds of treatment with hair dryer for 30min or baking at 70°C for 30min significantly reduced filterability of mask but one-round of treatment did not affect filterability of mask. Thus, we provided a simple method for reuse of disposable medical mask, which may help tackle the urgent need of medical mask during COVID-19 prevalence.

## Materials and Methods

### Cell and virus

Madin-Darby canine kidney (MDCK) cells were purchased from Cell Bank of the Chinese Academy of Sciences, Shanghai, China (www.cellbank.org.cn) and maintained in Dulbecco’s modified medium supplemented with 10% FBS (Gibco) and 25mM HEPES (Gibco) with non-essential amino acids (Gibco). Influenza A(H1N1) A/PR8 were purchase from ATCC and propagated in MDCK cells.

### Heat treatment of medical mask

Medical mask was mock-contaminated by dropping a 10μl of IVA solution containing 3.6×10^5^ pfu of IVA in 3 different sites on medical mask where each site was marked with a square. Due to the hydrophobic property of mask, liquid drop is difficult be absorbed onto the surface of the mask. The virus drop was doted on between the kink marks of the mask. Then the mask was wrapped with a food storage bag and then heat treated with hair dryer (1400w, 50Hz) by moving around over the surface of the wrapped mask vertically at a distance of about 10∼20cm. Alternatively, the wrapped mask was moved into baking oven and treated at indicated temperatures with various intervals.

### Rescue of the virus from mask

After heat treatment, the traces of IVA drop were visible within the marked squares. Due to the hydrophobic property of mask, the virus within the marked squares could be easily rescued by pipetting 250µl of Viral Transport Media (Hank’s Balanced Salt Solution (1X HBSS) supplemented with 0.1% bovine serum albumin (BSA), 100 units/ml penicillin G and 100 units/ml streptomycin)^9^ onto the traces until the traces disappear, then 125µl of the rescued virus was used for infecting naïve MDCK cells in the presence of 5µg/ml TPCK-trypsin (sigma) for 1 hour at 37°C in a cell culture incubator. After wash with DMEM media for 4 times, fresh DMEM media with 1µg/ml TPCK-trypsin was added. At indicated time points post infection, the supernatants were collected.

### Viral RNA quantification

The RNAs in the supernatants from the infected MDCK cells were extracted by QIAamp Viral RNA kit (Qiagen) according to the manufacturers’ protocol. RNAs were eluted in H_2_O and quantified by a Taqman-based real time PCR with One Step PrimeScript RT-PCR kit (Takara) according to the manufacturers’ protocol with 400nM of each primer (sense primer: CTT CTA ACC GAG GTC GAA ACG TA, anti-sense primer: GGT GAC AGG ATT GGT CTT GTC TTT A) and 200nM of probe (FAM-TCA GGC CCC CTC AAA GCC GAG-BHQ1). The relative RNA levels were calculated by 2^-ΔCT^ method ^10^.

### PM_2.5_ filterability measurement

A suction flask (Shuniu, 250mL, GG-17) together with the particulate matter monitor (TSI, AM510) were used to evaluate the PM_2.5_ filterability of medical masks (YINUO, FE3) before and after heat treatment. Considering the effect of suction process on the filtration performance of the masks, 5 experimental groups were set up as following:1). Untreated group (clean unused masks without any treatment); 2). One-round use + baking group (masks are used under normal condition for 4 hours and then baked in oven at 70°C for 30min); 3). Two-round use+ baking group (masks are used under normal condition for 8 hours, with incubation in oven at 70°C for 30min every 4 hours); 4). One-round use + hair dryer group (masks are used under normal condition for 4 hours and then heat treated by hair dryer for 30min as described above); 5). Two-round use + hair dryer group (masks are used under normal condition for 8 hours, with heat treatment by air dryer for 30min every 4 hours). Masks (*N*=3/group) were tightly bound to the top of the suction flask, while the particulate matter monitor was used to detect the PM_2.5_ concentrations near the top and the side outlet of the flask. The total suction filtration process lasted for 4 hours, and the tested flow rate is 1.8 L/min. PM_2.5_ concentrations were recorded every 30min, and repeated for 3 times at each time point.

## Results

### Heat inactivation of IVA on mock-contaminated medical mask

In our previous study, baking at 56°C for 30 min only partially inactivated IVA on medical mask, with a 1 ∼ 3 log reduction of viral RNA level, comparing to 6 log reduction of viral RNA level after hair dryer treatment for 30 min ^8^. In the present study, we first increased baking temperature and treated IVA mock-contaminated medical mask at 65°C for 30 min. After treatment, the virus on the mask was rescued and used to re-infect MDCK cells. Then we collected the supernatants of the infected cells at 0 and 48 hours post infection and quantified the viral RNA levels by real-time PCR. The inactivation efficiency was assessed by reduction of the viral RNA level comparing to the un-treated group. Baking at 65°C for 30 min resulted in 2 ∼ 5 log reduction of viral RNA level but did not reduce viral RNA level to basal level (level at 0 hour post infection) (Fig 1A). Hair dryer treatment for 30 min nearly completely inactive IVA on medical mask ^8^, while for convenience, we sought to shorten the treatment intervals. We assessed the inactivation efficiency of hair dryer treatment for 5, 10 and 15 min as described above. As shown in Figure 1A, hair dryer treatment for 5, 10 and 15 min did not effectively inactive IVA.

**Figure 1.**
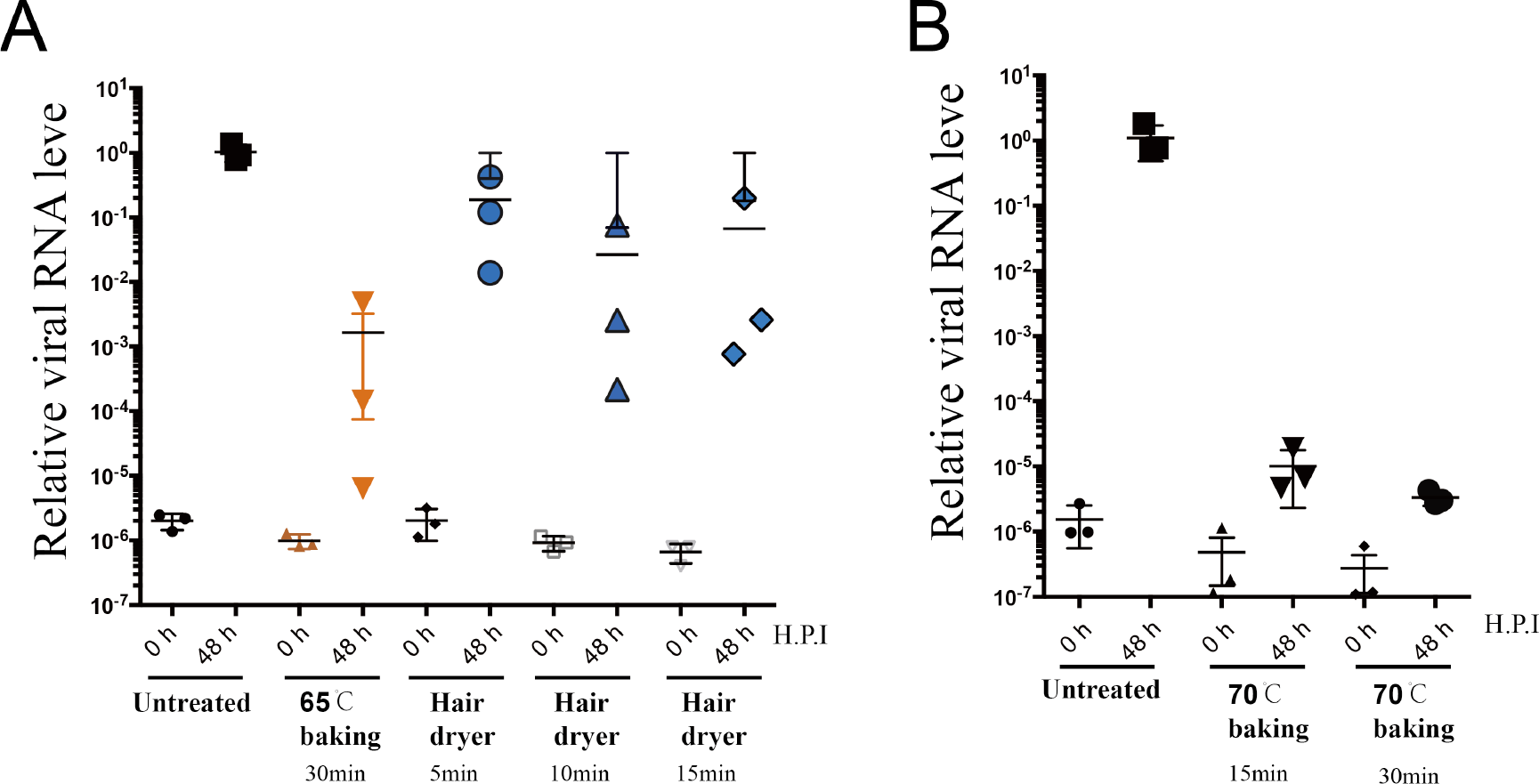
Assessment of efficiency of heat inactivation of IVA on mask. After indicated heat treatment, IVA on the mask was rescued and used to infect naïve MDCK cells. After extensive wash, the supernatants were collected at 0 and 48 hours post infection (H.P.I) and the virus in the supernatants were quantified by q-PCR. The RNA copy levels relative to that of the untreated (48 h) group were calculated and plotted. Mean ± SEM are shown (n=3).

Then we continued to optimize baking conditions and increase temperature to 70°C.We treated IVA-contaminated medical mask at 70°C for 15 min and 30 min and assessed the viral replication level of the rescued virus. As shown in Figure 1B, baking at 70°C for 15 min and 30 min both effectively reduced viral RNA levels to nearly basal levels and treatment for 30 min was more effective than treatment for 15min. Thus heat treatment with hair dryer for 30min ^8^ and baking at 70°C for 30 min both effectively inactive IVA-contaminated medical mask.

### PM_2.5_ filterability of medical masks after heat treatment

We assessed protective function of medical mask by measuring the filterability of medical masks to ambient PM_2.5_ after similar heat treatment as described before ^8^. To evaluate the recycle times of the medical mask, masks were subjected to several rounds of wear (use) and heat treatment. Masks were used under normal condition for 4 hours followed by treatment with hair dryer for 30min or baking at 70°C for 30 min, and then subjected to a 4-hour suction process mimicking the usage scenario by an adult under normal condition. As demonstrated in Figure 2A, PM_2.5_ filtration rate of all the tested disposable medical masks was slightly changed during the 4-hour suction process. In general, the filterability remained stable with PM_2.5_ filtration rate above 85% at all the tested time points. Among all the experimental groups, masks that have been used and heat treated once (by baking or hair dryer) have similar 4-hour average PM_2.5_ filtration rate compared to the untreated ones (Figure 2B). However, after being used and heat-treated twice (by baking or hair dryer), their filtration efficiency was significantly reduced as shown in Figure 2B. Our results indicate that heat inactivation by baking or hair dryer for 30 min may serve as an effective way for reuse of disposable medical masks, but this method is not encouraged to be used for more than once.

**Figure 2.**
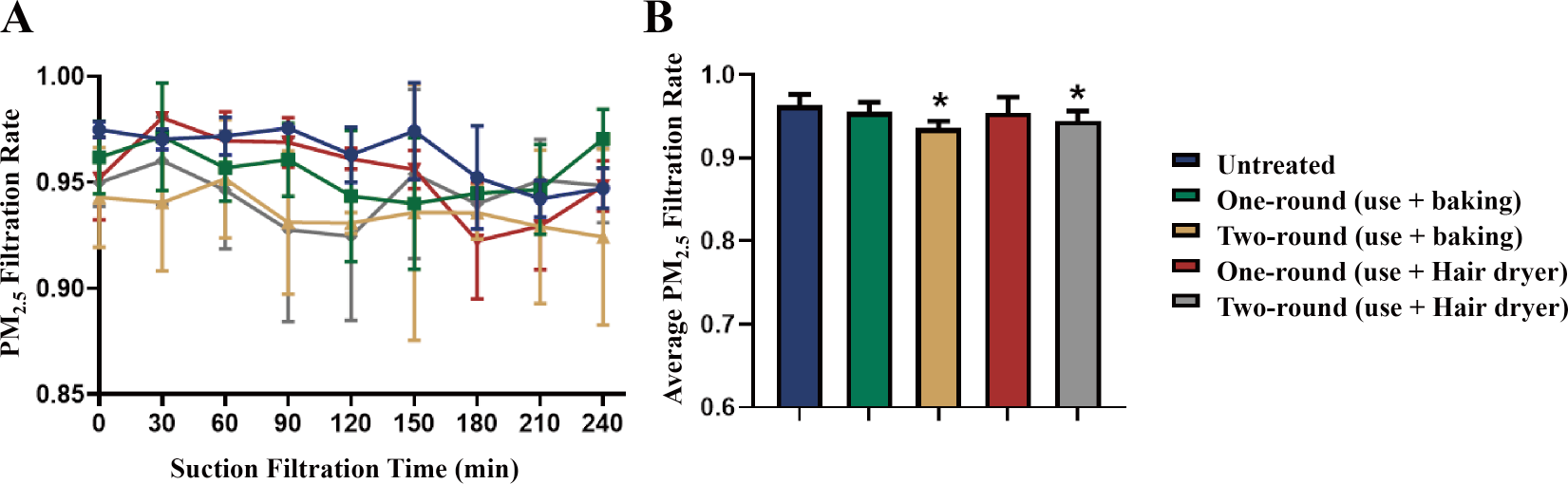
PM_2.5_ filterability of disposable medical masks. After one-round or two-round use and heat treatment, the filtration rate was measured. A. PM_2.5_ filtration rate change during the 4-hour suction process. \**p*<0.05 versus untreated group at the same time point, One-way ANOVA; B. Average PM_2.5_ filtration rate within the suction process were plotted. \**p*<0.05 versus untreated group, One-way ANOVA. Mean ± SD are shown (n=3).

## Discussion

It is not suggested to reuse disposable medical mask, especially for people who are in close contact with patients, medical staffs and laboratory technicians. But for community people who are at relative low risk of contacting high dosage of SARS-CoV-2, it is plausible to reuse medical mask after reliable inactivating treatment during the COVID-19 prevalence, which may alleviate the heavily global shortage of medical mask. We used IVA to mimic SARS-CoV-2 as they transmit similarly and both have envelopes and mock contaminated medical mask by dotting IVA drops onto the mask, then inactivated the IVA on medical mask by convenient heat treatment with hair dryer and baking that are easily to be adapted by community people. We found that baking at 70°C for 30min almost completely inactivated virus as hair dryer treatment for 30min described before ^8^. But baking at 56°C and 65°C for 30min could not effectively activate virus ^8^(Fig. 1A). Incubating at 56°C for 30min is a widely used protocol to inactivate influenza virus or coronavirus in solution. The incomplete inactivation by baking at this temperature and at 65°C indicates that baking a surface has less efficiency than incubating in solution. Maintaining the filter function of medical mask during usage is critical for preventing virus infection. We demonstrated that one-round of heat treatment did not affect filterability of mask whilst two-rounds of treatment slightly but significantly reduced filterability of mask. Thus, we provided a simple method for reuse of medical mask by heating with hair dryer for 30min or baking at 70°C for 30min and the treated mask can be reused at least once.

## Data Availability

The data is available to all the readers

## Acknowledgements

This work was inspired by professor Yuemei Wen. We are grateful to Chun-fai Yung and many other colleagues for their kind suggestions and inputs. This work was supported in part by the National Science and Technology Major Project of China (2017ZX10103009). Key Emergency Project of Shanghai Science and Technology Committee (20411950103). The funders had no role in study design, data collection and analysis, decision to publish, or preparation of the manuscript.

